# Screening of Thalassemia Carriers, Hemoglobin Variants, and Comparison of Hematological Parameters between Children of Bangladesh and Forcibly Displaced Myanmar Nationals

**DOI:** 10.1101/2023.09.19.23295805

**Authors:** Rumana Mahtarin, Kasrina Azad, Mohabbat Hossain, Mst. Sharmin Aktar Mukta, Mohammad Tanbir Habib, Abu Bakar Siddik, Nishat Sultana, Zannat Kawser, Umme Kulsum, Nusrat Sultana, Farjana Akther Noor, Ahmad Zubair Mahdi, Muhammad Asaduzzaman, Md. Ahashan Habib, Emran Kabir Chowdhury, Firdausi Qadri, Mst. Noorjahan Begum, A.H.M. Nurun Nabi

## Abstract

Thalassemia is a hereditary blood disorder with different severity spectrums. This study aimed to assess thalassemia screening rates between children of Bangladesh and selected camps of Forcibly Displaced Myanmar Nationals (FDMN) in Cox’s Bazar in Bangladesh and compare the hematological parameters among the screening groups. Complete blood count (CBC) analysis and hemoglobin electrophoresis for each participant were performed by collecting venous blood. Statistical analysis was employed for the comparison of parameters in blood. The thalassemia carrier and other hemoglobin variant rate in Bangladeshi children in selected areas have been found to be 20.7% and in FDMN children, the rate is 8.2%. Hematological differences are visualized among children of two nations. Intra-and interpopulation variances are highlighted in principal component analysis where higher variance (94.87%) in Bangladeshi children than FDMN children (80.68%). Receiver operating characteristics (ROC) and area under the curve (AUC) analyses revealed, RBC (0.761, 0.902, and 0.791) and RDW-CV are better model (0.819, 0.925, and 0.858) among the classifier of blood parameters. Pearson correlation shows distinguished covariation or association among the parameters. The outcome of the study highlights the discrepancies in levels of carriers in regions in Bangladesh and suggests further screening as well as population based molecular research to ensure better treatment strategies.

## 1. Introduction

Thalassemia is an inherited hemolytic anemia which passes through autosomal recessive pattern [1]. A fractional or complete absence in the synthesis of α-globin chains or β-globin chains due to genetic defects in the *α-globin* gene cluster on chromosome 16 or the *β-globin* gene cluster on chromosome 11 can lead to the synthesis of defective hemoglobin [2]. In β-thalassemia, the main consequence of unstable hemoglobin and iron overload is oxidative stress [4]. Iron triggers the manufacture of reactive oxygen species, which in excess are toxic, contributing to oxidative damage to red blood cells, leukocytes (recurrent infections), platelets (hypercoagulable state) [4], cellular dysfunction, apoptosis, and necrosis in vital organs such as heart, liver and endocrine system [1, 2]. People with β-thalassemia major (β 0 /β0, β0 /β+, and sometimes β+ / β+) generally get medical attention within the first two years of life and necessitate regular RBC transfusions to survive [2]. This transfusion can cause iron overload [3]. Dysregulation of iron homeostasis promotes iron deposition in the organs and influences disease severity, which is the main cause of death in beta-thalassemia patients [4, 5]. Each year worldwide more than 300,000 newborns are detected with major hemoglobinopathies of whom 60,000 to 70,000 are β-thalassemia major cases, especially in the Mediterranean area, Middle East, Far East, and East Asia [3]. In Bangladesh, the birth rate of 33/10,000 had been reported where 8.68% had HbE trait, 2.24% had β-thalassemia (β-thal) trait and nearly 2500 thalassemia major cases are added every year in Bangladesh [6,7] while in Myanmar, thalassemia carrier rate was 4.3% [8]. Previously migration increased the frequency of thalassemia disease in lower-prevalence regions [10]. Over a million Rohingyas from Myanmar were forcefully displaced to Bangladesh and other neighboring countries by January 2018. Over half of the displaced Rohingyas have sought shelter in Bangladesh and settled in Rohingya camps in Cox’s Bazar [11]. There are no published thalassemia screening outcomes on Forcefully Displaced Myanmar Nationals (FDMN) in Bangladesh. This study aimed to compare the prevalence of thalassemia carriers and hemoglobin variants to associate hematological parameters among Bangladeshi children and Rohingya children who were forcibly displaced from Myanmar in Bangladesh.

## 2. Methods

### 2.1. Enrollment of Study Participants

In screening campaigns, a total of 92 Bangladeshi children (49% male and 51% female), aged between 1 and 15 years and 110 children of forcibly displaced Myanmar Nationals (58% male and 42% female), aged between 7 and 16 years were enrolled in the study from 2 May 2022 to 22 October 2022. The study (# PNR-22003) was ethically approved by Institutional Review Board of Institute for Developing Science and Health Initiatives (ideSHi). Participants were provided informed consent and written consent was taken from parents or legal guardians of each participant.

### Sample size calculation

Required sample size has been calculated by the following formula [12, 13]

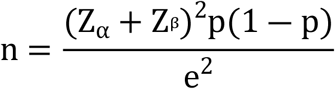

Where,

n = required sample size,

*Z*_1*―α* 2_= Standard normal variate for level of significance

Z_*β*_= Standard normal variate for power

p = Expected prevalence,

e = Margin of error or acceptable difference

n_b_ = Sample size for Bangladeshi Population.

n_m_ = Sample size for Myanmar Population.

Considering, the level of significance = 5%, power of the test = 85%, expected prevalence rate for Bangladeshi population P_b_ =10.2% [14], margin of error e=10% and including 15% attrition rate the sample size is n_b_ = 92. Again, with the level of significance = 5%, power of the test = 96%, expected prevalence rate for Myanmar population P_m_ =4·3% [8], margin of error e=9% and including 15% attrition rate the sample size is n_m_ = 110.

### 2.2. Analysis of Complete Blood Count of The Study Participants

Three ml of venous blood was collected from each participant in EDTA containing vacutainer and the collected blood samples were transported to the laboratory maintaining 4-8°C and preserved at 4°C until analysis. The complete blood count (CBC) was performed for each sample by using XS-800i Hematology Analyzer (Sysmex, Japan) following the manufacturers’ instruction. Major hematological parameters like hemoglobin (HGB), hematocrit (HCT), RBC count and Red Cell Indices including mean corpuscular volume (MCV), mean corpuscular hemoglobin (MCH), mean corpuscular hemoglobin concentration (MCHC) and Red cell distribution width (RDW) were considered for analysis.

### 2.3. Hemoglobin Electrophoresis

Hemoglobin electrophoresis was performed on automated capillary electrophoresis system (Sebia, France) using Capillary Hemoglobin (E) kit to measure HbA, HbA2, HbF, and other abnormal Hb variants following manufacturer’s instructions.

### 2.4 Statistical Analysis

Statistical analysis was performed using SPSS and GraphPad Prism software (version 9.0) and R programming. P < 0.05 was considered statistically significant for all the tests. One-way ANOVA was performed for the analysis of hematological differences, and the distribution pattern was visualized. Principle component analysis was performed for understanding intra and inter population variation. Covariation of association among the major hematological parameters was visualized by Pearson correlation. Receiver operating characteristics (ROC) and area under the curve (AUC) analyses were also conducted for diagnostic assessment.

## 3. Results

### 3.1. Thalassemia Carrier and Hemoglobin Variant Screening Outcomes

In the study, the screening showed that the frequency of thalassemia traits and other hemoglobin variants in Bangladeshi children was 20.7%, where beta thalassemia trait (BTT) and HbE trait (HbET) were 5.43% and 13.04%, respectively. The highest carrier frequency was found among the children in Dhaka and Rangpur (only HbE, 23.5%) divisions, while it is comparatively lower in Chattogram (17.65%, BTT-11.76%, HbET-5.89%), and Barishal (17.65%, BTT-11.76%, HbET-5.89%). The lowest carrier frequency was observed among the children in Sylhet (only HbET-5.8%), Mymensingh (only HbET-5.8%) and Khulna (only BTT-5.8%) divisions. The low HbA2 had been detected in Rajshahi (1.1%).

On the other hand, thalassemia carrier and other hemoglobin variant frequency was 8.2% among the children from the FDMNs, where BTT and HbE were 2.73% and 5.45%, respectively. In addition, HbD variant was also found in the Bangladeshi children (1.09%) and FDMN children (0.9%). The distribution of different groups in screening was displayed in Fig 1.

**Fig 1.**
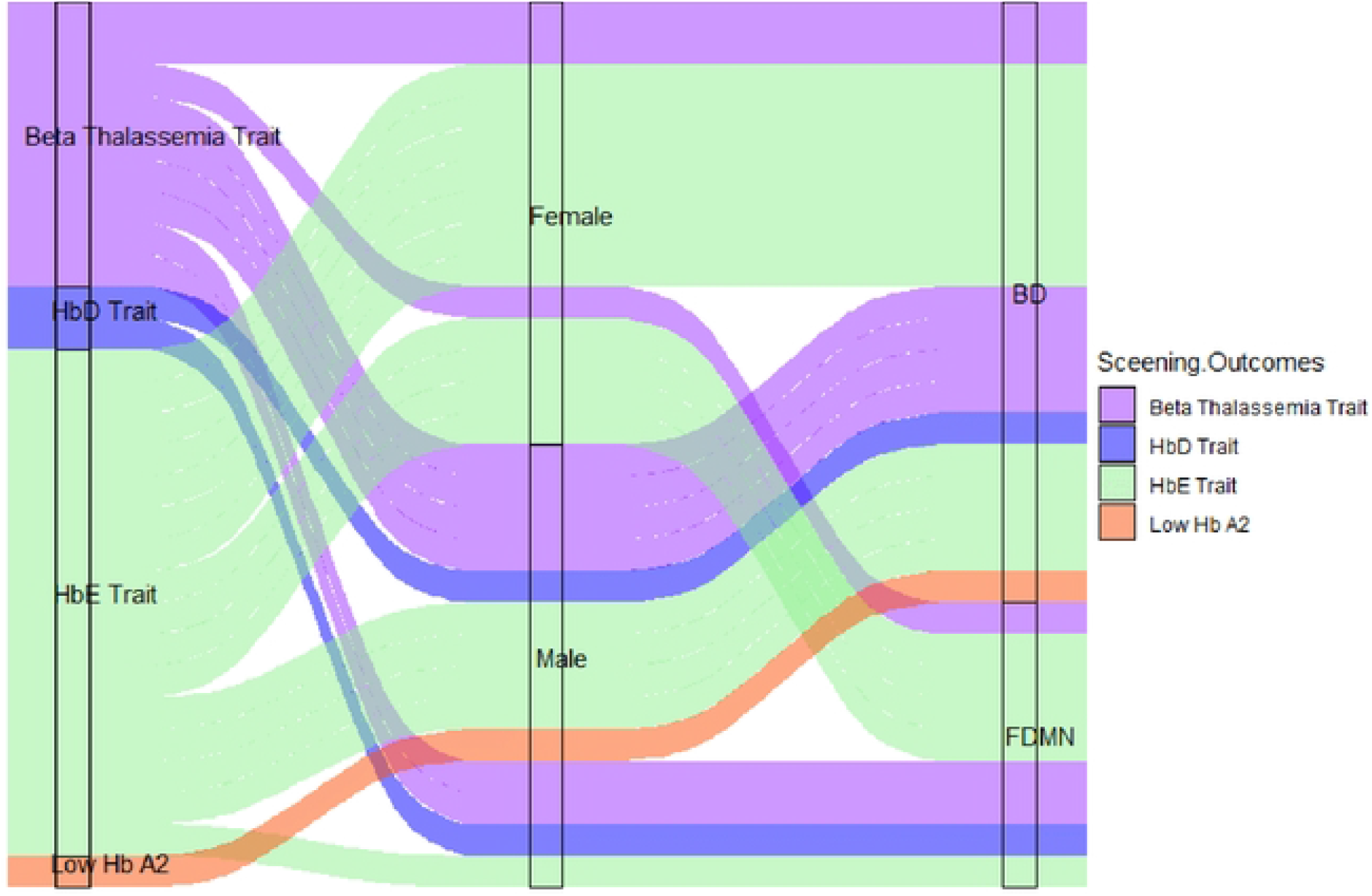
The alluvial plot represents distribution of screening outcomes in BD and FDMN children The color codes indicate different groups: Beta Thalassemia Trait (purple), HbD Trait (blue), HbE Trait (lemon), Low Hb A2 (orange).

### 3.2. Analysis of Hemoglobin Variants

Hemoglobin electrophoresis among the healthy participants of Bangladeshi children showed the levels of HbA and HbA2 were 97.14 ± 0.31% and 2.80 ± 0.25%, respectively. In beta thalassemia trait (BTT), the levels of HbA, HbA2 and HbF were 93.78 ± 1.5%, 5.40 ± 0.36%, and 2.05 ± 2.33% respectively. In HbE trait the levels of HbA, HbA2 and HbE were 70.80± 1.4%, 3.72 ± 0.48%, and 25.39± 1.08%, respectively. In addition, HbA (54%), HbA2 (3.7%), and HbD (42.3%) were found in HbD trait. In low HbA2 participant, HbA was 98.1% and HbA2 was 1.9%, and suspected to have alpha thalassemia trait (HGB 8.7 g/dL, RBC 4.68 X10^12/L, MCV 55.8 fL, MCH 18.6 pg) according to Mentzer index (MCV/RBC <13).

On the other hand, among the normal or non-carrier children of FDMN, the levels of HbA and HbA2 were 97.3 ± 0.29% and 2.67 ± 0.21% respectively. In beta thalassemia trait (BTT), the level of HbA was 93.6 ± 0.47%, HbA2 was 5.3 ± 0.47 % and HbF was 1.17 ± 0.5%. In HbE trait, the level of HbA was 72.9 ± 3.12%, HbA2 was 3.8 ± 0.35%, HbE was 23.0± 3.4%, and HbF was 1.1%. In addition, a child of FDMN with HbD trait showed the level of HbA, HbA2, and HbD at 56.4%, 2.9%, and 41%, respectively. Alpha thalassemia trait was not found in FDMN children from study population.

### 3.3. Principal Component Analysis

In this study, the principal component analysis was considered as feature extractor to compare the participants of Bangladesh (BD) and Forcibly Displaced Children of Myanmar Nationals. The principal components, PC1 and PC2 accounted for most of the variation in the participants based on gender and age distribution. In Fig 2A among Bangladeshi children, PC1 and PC2 showed 63.0% and 31.7% variance, respectively (in total 94.87%). On the contrary, the FDMN children showed the variance of PC1 and PC2 at 50% and 30.7%, respectively, while the total variance was 80.68% (Fig 2B).

**Fig 2.**
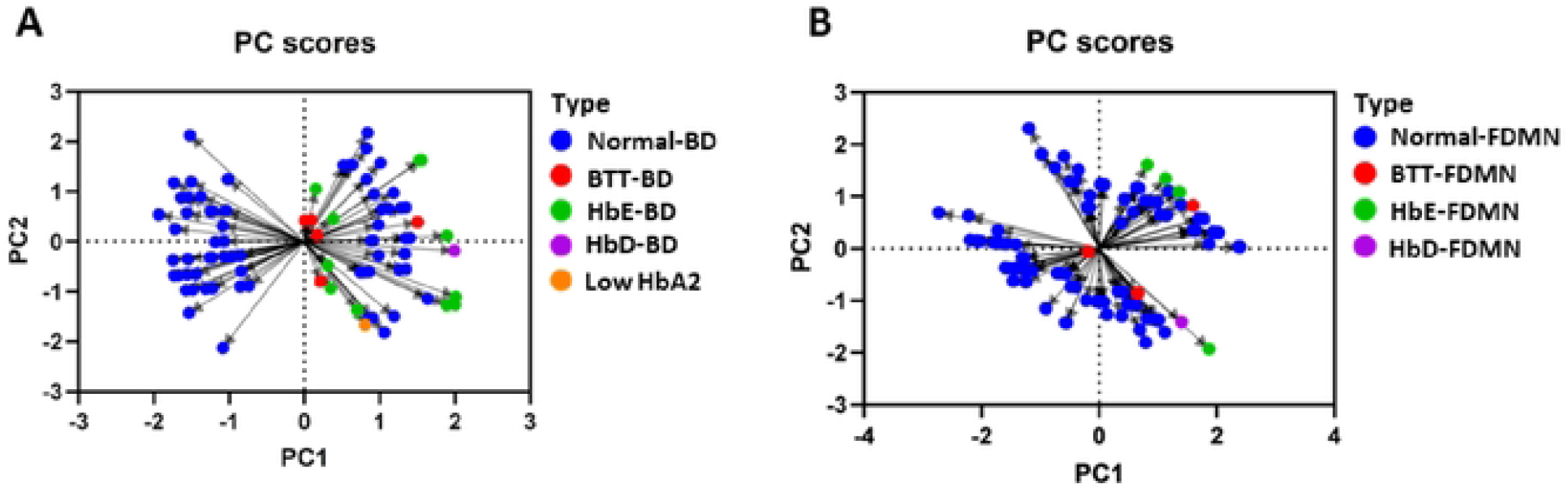
Intra-and inter-population variances are highlighted in Principal component analysis. (A) PC scores in BD children’s groups and (B) PC scores in FDMN children’s groups. The color codes represent different groups in BD and FDMN children, Normal (blue), Beta Thalassemia Trait (BTT) (red), HbE (green), HbD (purple), Low HbA2 (orange).

### 3.4. Analysis of Hematological Differences between **Bangladeshi** and Forcibly Displaced Children of Myanmar Nationals

The column bar graphs exhibited a diverse distribution of major hematological parameters among the prevalent groups in children of the two countries.

According to the one-way ANOVA test, RBC, MCV, MCH, and RDW-CV were substantially different among the normal, BTT, and HbET groups in Bangladeshi children (Fig 3A), with a p-value of 0.05. Among the BTT children of Bangladesh, RBC and RDW-CV were found significantly higher, whereas MCV and MCH were significantly lower compared to the normal value (Table 1). In the FDMN children, all the hematological parameters except MCHC showed significant results among the normal, BTT and HbET groups (Fig 3B) with a p-value of <0.05. RBC and RDW-CV were significantly higher, while HGB, HCT, MCV, and MCH were significantly lower compared to the normal level in BTT in the FDMN children (Table 2).

**Table 1.** Comparative analysis of major hematological parameters of Bangladeshi children in major groups.

| Hematological parameters | Normal | BTT | HbET | P value |
| --- | --- | --- | --- | --- |
| RBC (X10<sup>12</sup>/L) | 4.34 ± 0.47 | 5.83 ± 1.14 | 4.76 ± 0.50 | 0.000 |
| HGB (g/dL) | 11.53 ± 1.25 | 10.87 ± 1.68 | 10.79 ± 1.21 | 0.107 |
| HCT (%) | 31.59 ± 3.27 | 30.50 ± 4.55 | 29.56 ± 2.59 | 0.096 |
| MCV (fL) | 72.97 ± 4.98 | 52.73 ± 3.54 | 62.61 ± 6.06 | 0.000 |
| MCH (pg) | 26.62 ± 1.89 | 18.77 ± 1.17 | 22.68 ± 2.46 | 0.000 |
| MCHC (g/dL) | 36.49 ± 1.01 | 35.60 ± 0.26 | 36.19 ± 1.21 | 0.232 |
| RDW-CV (%) | 13.52 ± 1.01 | 17.73 ± 1.94 | 15.19 ± 2.24 | 0.000 |

Here, RBC means Red Blood Corpuscle, HGB means Hemoglobin, HCT means Hematocrit, MCV means Mean Corpuscular Volume, MCH means Mean Corpuscular Hemoglobin, MCHC means Mean Corpuscular Hemoglobin Concentration, and RDW-CV means Red Cell Distribution Width - Coefficient of Variation

**Table 2.** Comparative analysis of major hematological parameters of Forcibly Displaced Children of Myanmar Nationals (FDMN) children in major groups.

| Hematological parameters | Normal | BTT | HbET | P value |
| --- | --- | --- | --- | --- |
| RBC (X10<sup>12</sup>/L) | 4.46 ± 0.33 | 5.45 ± 0.17 | 4.89 ± 0.26 | 0.000 |
| HGB (g/dL) | 11.41 ± 0.90 | 9.43 ± 0.65 | 10.90 ± 0.67 | 0.001 |
| MCV (fL) | 73.92 ± 4.03 | 51.03 ± 0.81 | 63.76 ± 3.85 | 0.000 |
| HCT (%) | 32.92 ± 2.39 | 27.83 ± 1.03 | 31.12 ± 1.70 | 0.001 |
| MCH (pg) | 25.62 ± 1.71 | 17.30 ± 0.66 | 22.34 ± 1.45 | 0.000 |
| MCHC (g/dL) | 34.64 ± 0.99 | 33.90 ± 1.15 | 35.02 ± 0.66 | 0.299 |
| RDW-CV (%) | 13.40 ± 0.80 | 19.50 ± 0.95 | 14.32 ± 0.30 | 0.000 |

Here, RBC means Red Blood Corpuscle, HGB means Hemoglobin, HCT means Hematocrit, MCV means Mean Corpuscular Volume, MCH means Mean Corpuscular Hemoglobin, MCHC means Mean Corpuscular Hemoglobin Concentration, and RDW-CV means Red Cell Distribution Width - Coefficient of Variation

**Fig 3.**
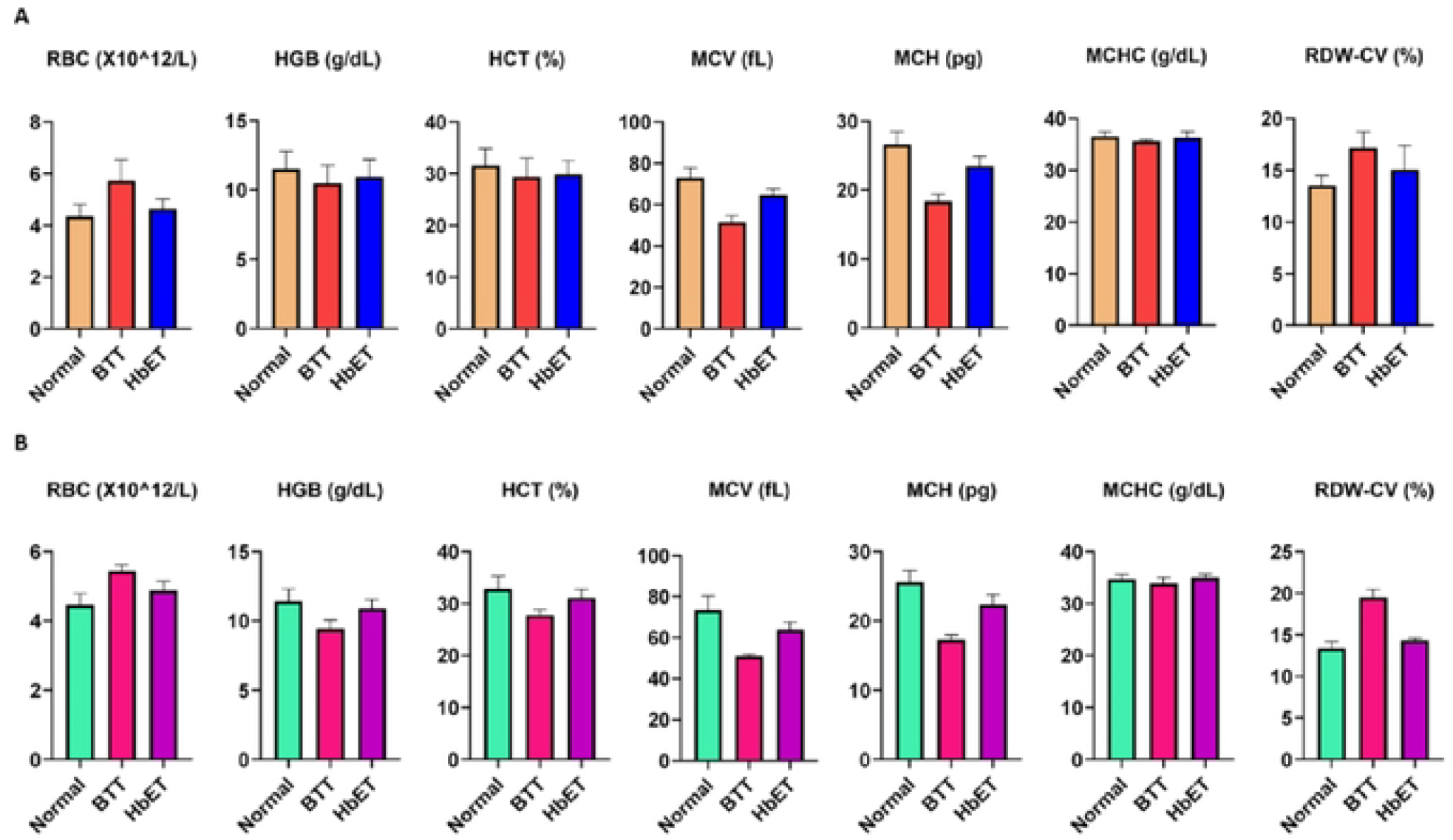
Distribution pattern of major hematological parameters. Normal, Beta Thalassemia Trait (BTT), and Hemoglobin E Trait (HbET) groups in both (A) BO and (B) FDMN children.

However, the hematological parameters of both Bangladeshi children and FDMN children showed significant results for RBC, HGB, MCV, MCH, HCT, and RDW-CV among the normal, BTT and HbET groups. RBC and RDW-CV were significantly higher, while HGB, HCT, and MCH were significantly lower in BTT of both populations (Table 3).

**Table 3.** Comparative analysis of major hematological parameters of both Bangladeshi and Forcibly Displaced Children of Myanmar Nationals (FDMN) children in major groups Hematological parameters Normal BTT HbET P value.

| Hematological parameters | Normal | BTT | HbET | P value |
| --- | --- | --- | --- | --- |
| RBC (X10<sup>12</sup>/L) | 4.41 ± 0.40 | 5.62 ± 0.64 | 4.71 ± 0.37 | 0.000 |
| HGB (g/dL) | 11.46 ± 1.06 | 10.09 ± 1.18 | 10.93 ± 1.09 | 0.000 |
| MCV (fL) | 72.98 ± 5.36 | 58.51 ± 9.04 | 66.58 ± 5.44 | 0.000 |
| HCT (%) | 32.36 ± 2.90 | 29.80 ± 3.04 | 30.79 ± 2.85 | 0.013 |
| MCH (pg) | 26.04 ± 1.85 | 17.96 ± 1.01 | 23.16 ± 1.47 | 0.000 |
| MCHC (g/dL) | 35.42 ± 1.35 | 34.96 ± 1.10 | 35.91 ± 1.26 | 0.206 |
| RDW-CV (%) | 13.45 ± 0.89 | 18.03 ± 1.80 | 14.81 ± 2.01 | 0.000 |

Here, RBC means Red Blood Corpuscle, HGB means Hemoglobin, HCT means Hematocrit, MCV means Mean Corpuscular Volume, MCH means Mean Corpuscular Hemoglobin, MCHC means Mean Corpuscular Hemoglobin Concentration, and RDW-CV means Red Cell Distribution Width - Coefficient of Variation

### 3.5. Correlation among Hematological Parameters in Different Groups of Children in Bangladesh and Forcibly Displaced Myanmar Nationals (FDMN)

Pearson correlation showed covariation or association among the hematological parameters. Diverse correlation pattern in CBC parameters was persistent in both BD and FDMN children. Among Bangladeshi children, normal or non-carriers and HbET showed quite similar type of strong positive correlation pattern among RBC, HGB, HCT, MCV and MCH, while RDW-CV exhibited negative correlation with the other hematological parameters. BTT showed strong positive relationship among RBC, HGB, HCT, and RDW-CV, MCV, and MCH. In FDMN, variations were more prominent among the different groups. BTT exhibited strong positive correlation among most of the hematological parameters except HCT and RDW-CV (Fig 4).

**Fig 4.**
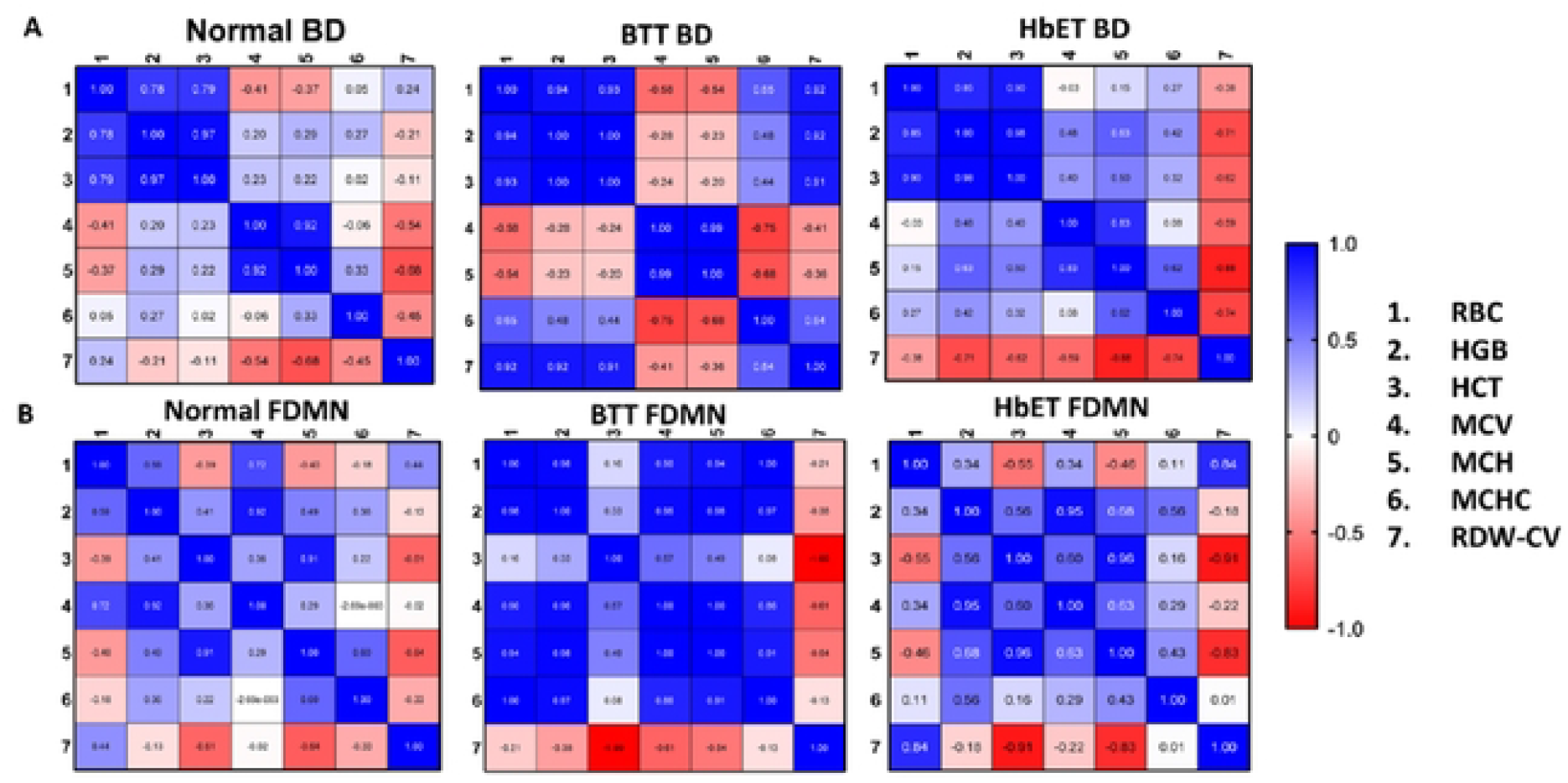
Correlation pattern in major hematological parameters in both BD and FDMN children in major groups. The groups in (A) BD children and (B) FDMN children. BTT indicates Beta Thalassemia Trait and HbET indicates Hemoglobin E Trait. The major hematological parameters are numbered as 1 to 7 and the correlation is measured as -1.0 to 1.0. Color code indicates correlation pattern. Red color indicates negative correlation (-1.0 to 0) where -1.0 indicates least correlation and blue color indicates positive correlation (0 to 1.0) where 1.0 indicates highest correlation.

### 3.6. ROC Curve Analysis

ROC curve analysis was performed for the assessment of diagnostic value of thalassemia carrier and hemoglobin variant screening. ROC curves were plotted for children of Bangladesh and FDMN children (Fig 5A, Fig 5B) and for both nations (Fig 5C). AUC was measured to visualize the classifier of blood parameter model. RBC (0.761, 0.902, and 0.791) and RDW-CV (0.819, 0.925, and 0.858) covered high AUC and good classifier for carrier screening. High AUC indicated more accuracy for diagnostic performance which might be supportive to hemoglobin electrophoresis test for proper diagnosis of thalassemia and hemoglobinopathies.

**Fig 5.**
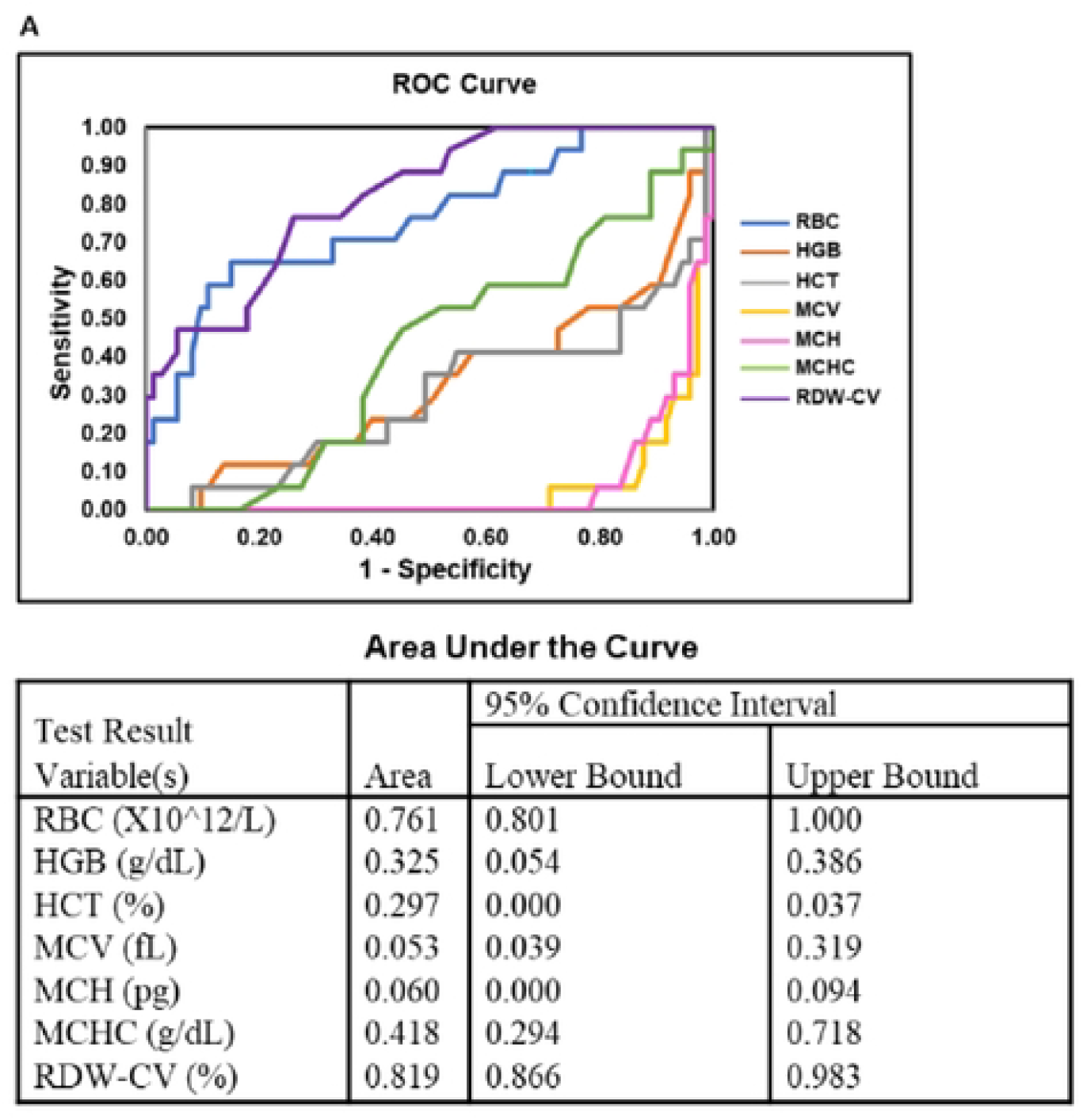
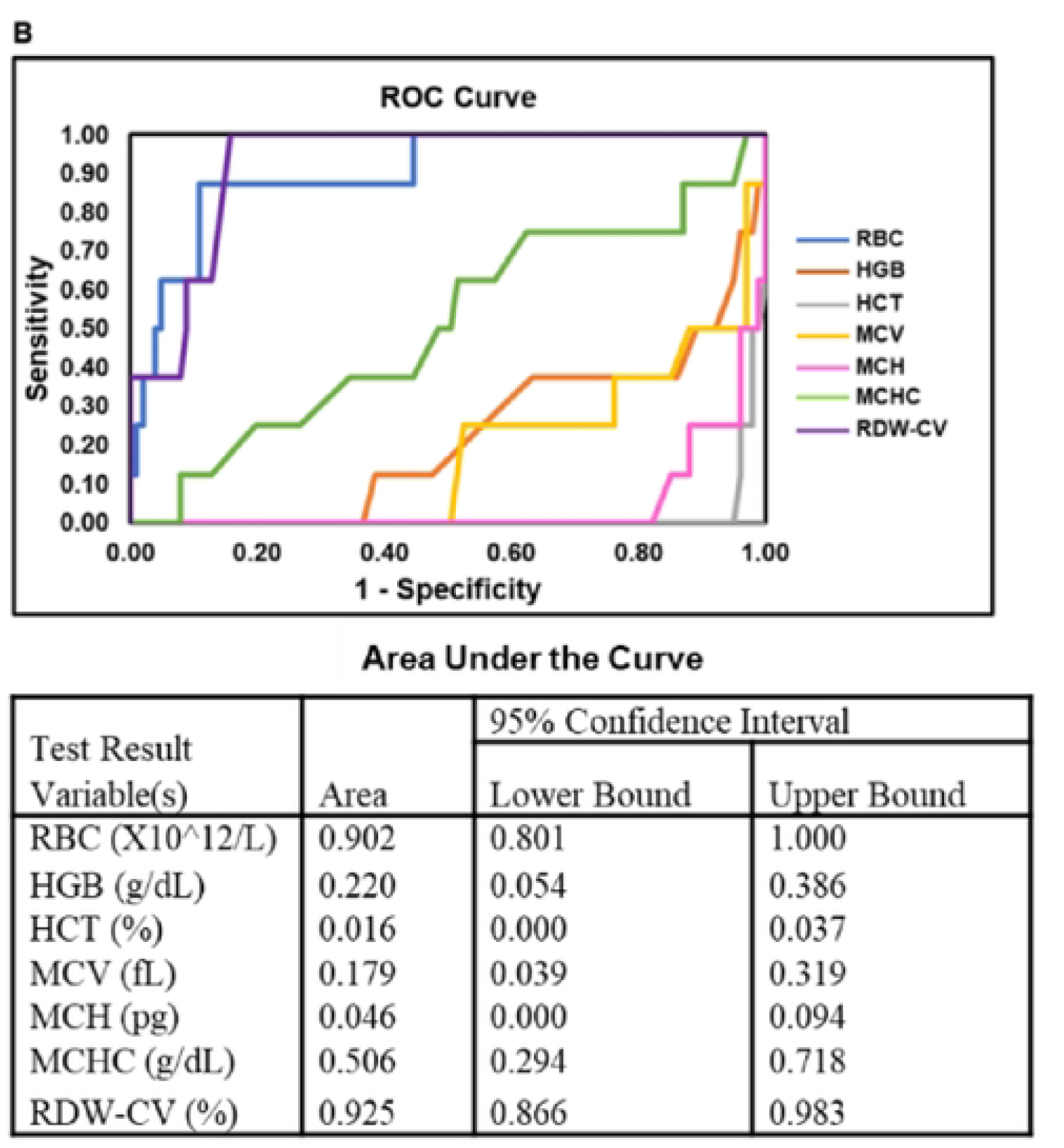
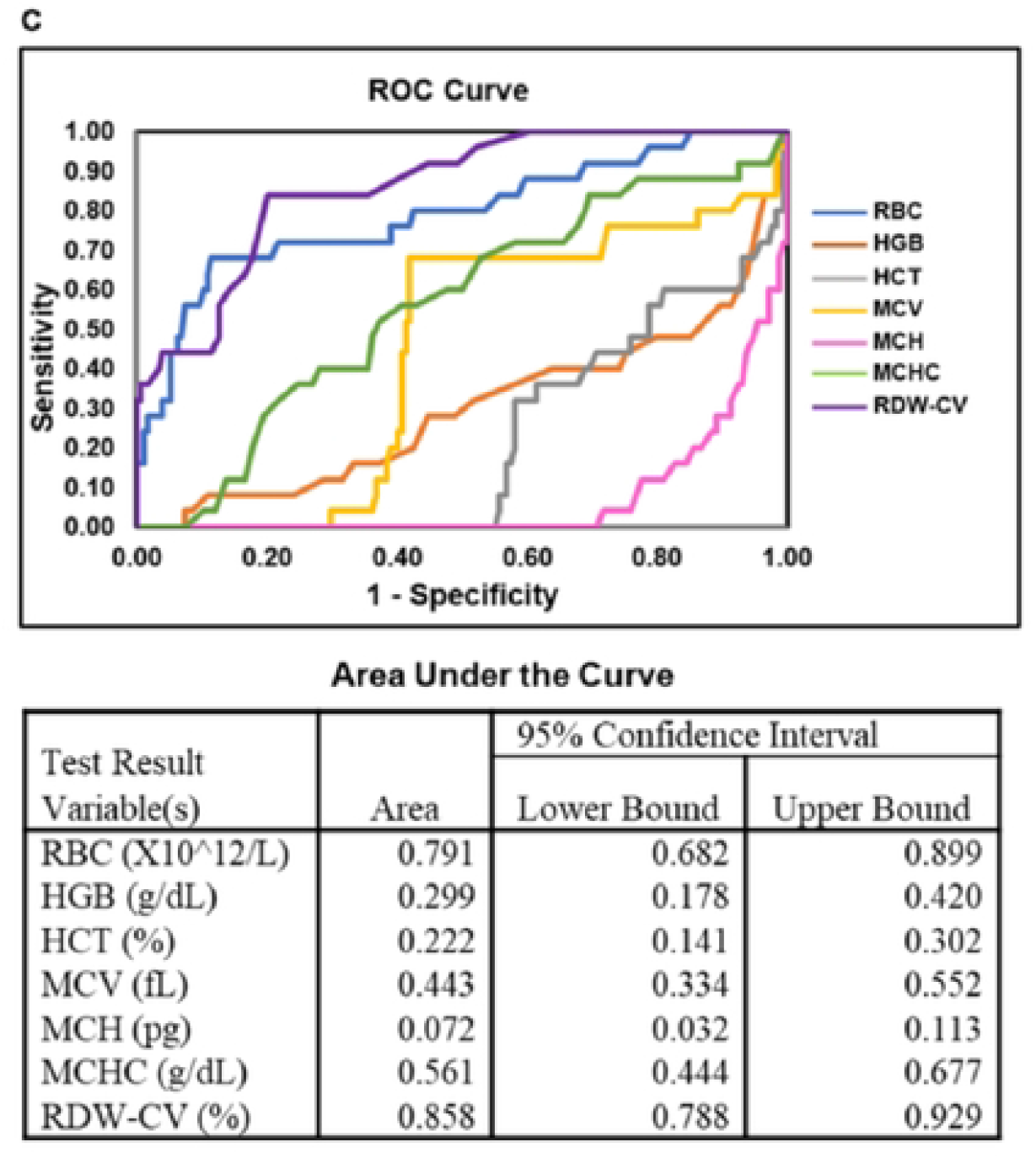
ROC curve and area under curve display classifier of blood parameter model. (A) BD (B) FDMN (C) both BD and FDMN.

## 4. Discussion

This is the first study for assessment of thalassemia carrier and hemoglobin variant screening data in children between Bangladesh and Forcibly Displaced Myanmar Nationals (FDMN). In the study, the frequency of thalassemia carriers and hemoglobin variants in Bangladeshi children is found 20.7%, which is double of the previous report (10.2%) [7, 14]. The highest carrier frequency was found in Dhaka and Rangpur divisions while the lowest carrier frequency was observed in Sylhet, Mymensingh, and Khulna divisions. On the other hand, the carrier rate in FDMN children was 8.2%, where beta thalassemia trait (BTT) rate is 2.73%, and HbE trait (HbET) rate is 5.45%. Combining the rate for both study groups, the cumulative carrier rate is around 14% which is also demonstrate the increasing trend compared to previous record [7, 14]. Principle component analysis highlights the marked variations in Bangladesh and Forcibly Displaced Myanmar Nationals for their genetic ancestry conformation. Higher variance has been exposed in Bangladeshi children (94.9%) than in FDMN children (80.7%) for intra-and inter-population variances of different groups. Divergent distribution pattern of major hematological parameters was visualized for the children of both Bangladesh and FDMN. The variations were higher in BTT group of both populations while wider distribution in Bangladesh and narrower distribution patterns are visible in FDMN children. ROC curve evaluates the diagnostic performance of carrier and hemoglobin variant screening by demonstrating the sensitivity and specificity of the test. AUC under ROC curve was measured to visualize the classifier of blood parameter model. RBC (0.761, 0.902, and 0.791) and RDW-CV (0.819, 0.925, and 0.858) display high AUC which indicates better classifier for the screening. Comparing all the parameters, RBC and RDW-CV are supportive to hemoglobin electrophoresis to evaluate the screening outcomes. There was association of RDW-CV and thalassemia disease severity in previous study [9]. Correlation matrixes represent covariation or association among the parameters. Distinguished correlation was revealed in both Bangladesh and FDMN children. BTT groups of both Bangladesh and FDMN demonstrate the higher number of strong positive correlation among parameters although the parameters differ in their association. Mostly RDW-CV demonstrates quite negative association with other hematological parameters in the FDMN children. All the analyses demonstrate the similarities and discrepancies in the major hematological parameters of the groups from Bangladesh and Forcibly Displaced Myanmar Nationals. The screening in children has been conducted to determine the carrier and hemoglobin variant rates of existing populations with two different genetic background in Bangladesh. However, a drawback is that we could screen only a limited number of children since this was conducted within a thalassemia awareness program. More extensive thalassemia carrier screening and awareness programs, prenatal diagnosis, and compulsory carrier screening before marriage should be implemented to prevent the spread of thalassemia disease and other hemoglobinopathies. This comparative study will be helpful for track the rates of thalassemia carrier and hemoglobin variant in children to prevent the disease burden. As well as this population-based study will expedite molecular analysis for the improvement of thalassemia treatment strategies by initiating personalized medicine. Finally, such studies and programs will help in creating awareness of thalassemia, its prevention and in policy changes in the country.

## 5. Conclusion

The frequency of carrier and hemoglobin variant is much higher in Bangladeshi children than the displaced population from Myanmar (FDMN) into Bangladesh. This comparative study highlights the discrepancies in thalassemia carrier and hemoglobin variant status on children which will be helpful to understand the burden of thalassemia and hemoglobinopathies. Increasing the number of screening and being aware of devastating health consequences are vital for prevention of disease.

## Data Availability

All relevant data are within the manuscript

## Acknowledgement

The authors are grateful to the First Security Islami Bank, Bangladesh for their contribution and support to conduct the thalassemia screening program. We are thankful to the study participants, the clinicians and staff for their assistance in specimen collection, and the thalassemia screening team members of ideSHi.

## Notes

### Competing Interest Statement

The authors have declared no competing interest.

### Funding Statement

Kasrina Azad received funds from First Security Islami Bank, Bangladesh (https://fsiblbd.com/). The funders had no role in study design, data collection and analysis, decision to publish, or preparation of the manuscript.

### Author Declarations

APPROVAL LETTER IRB of ideSHi PNR-22003 18 April 2022 Ms Rumana Mahtarin Principal Investigator of research protocol # PNR-22003, and PhD Fellow Institute for Developing Science and Health Initiatives (ideSHi) Sub: Approval of research protocol # PNR-22003 Approval Date: 18 April 2022 Review Type: Full Committee Review Risk Level: No more than minimal Project type: New Project Dear Ms Mahtarin, Thank you for your memo dated 15 April 2022 attaching the modified version of your research protocol # PNR-22003, titled “Molecular analysis of β-globin and iron regulatory genes in thalassemia carriers and different disease spectrums of HbE/β and β-thalassemia major patients in Bangladesh”, addressing the issues raised by the Institutional Review Board (IRB) in its March meeting held on 19 March 2022 at the ideSHi Meeting Room, to satisfaction of the Board. I am pleased to inform you that your protocol is approved.

